# Normative characterization of age-related periodic and aperiodic activity in resting-state clinical EEG recordings

**DOI:** 10.1101/2024.06.18.24308910

**Authors:** Sophie Leroy, Viktor Bublitz, Falk von Dincklage, Daria Antonenko, Robert Fleischmann

**Affiliations:** Delirium Prevention Unit, Universitätsmedizin Greifswald, Fleischmannstraße 6, 17489 Greifswald, Germany; Department of Neurology, Universitätsmedizin Greifswald, Fleischmannstraße 6, 17489 Greifswald, Germany; Department of Anesthesiology and Operative Intensive Care Medicine, Charité University Medicine Berlin, Campus Charité Mitte and Virchow-Klinikum, Augustenburger Platz 1, 13353 Berlin, Germany; Department of Anesthesia, Intensive Care, Emergency and Pain Medicine, Universitätsmedizin Greifswald, Fleischmannstraße 6, 17489 Greifswald, Germany

**Keywords:** EEG, brain oscillations, aging, periodic activity, aperiodic activity, tACS

## Abstract

**Objective:** Non-invasive oscillatory brain stimulation techniques target the oscillatory activity of the brain and have several potential therapeutic applications in the clinical context. The aim of these analyses from clinical routine EEG recordings was to provide normative values of physiological age-related oscillatory (periodic) and non-rhythmic (aperiodic) activity.

**Methods:** We analyzed 532 EEGs of patients between 8 and 92 years of age. EEG segments were preprocessed, and the power spectrum was computed using a multitaper method. We decomposed the power spectrum into periodic (peak power, frequency, and bandwidth) and aperiodic (intercept and exponent) components. Linear regression models were used to investigate age-related changes in these parameters.

**Results:** We observed significant global age-related changes in the periodic alpha (- 0.015 Hz/year) and gamma (+ 0.013 to + 0.031Hz/year) peak frequency as well as in the aperiodic exponent (- 0.003 to - 0.004 μV^2^/Hz/year). In the other parameters there were solely regional or no significant age-related changes.

**Conclusions:** Decomposing the power spectrum into periodic and aperiodic components allows for the characterization of age-related changes.

**Significance:** This study provides the first spectrum-wide normative characterization of age-related changes in periodic and aperiodic activity, relevant for non-invasive brain stimulation with alternating current targeting ongoing oscillatory activity.

**Highlights:**

- Alpha peak frequency decreases with age, while gamma peak frequency accelerates.
- Age-related changes in alpha and theta power result from a flattening of the aperiodic slope, not decreased oscillatory activity
- Decomposing the EEG spectrum into periodic and aperiodic activity is essential when characterizing ongoing oscillatory activity

## 1. Introduction

Electroencephalography (EEG) provides essential insights into the dynamic changes in neural activity across various life stages. The nuanced age-related alterations in both periodic and aperiodic EEG activity reveal complex patterns of neural development and aging (Donoghue et al., 2020). From early childhood to elderly adulthood, the aperiodic activity diminishes (Hill et al., 2022). These alterations may arise from a shift in the balance between oscillatory coupling and local population spiking (Voytek and Knight, 2015). Likewise, aging alters dynamic network communication, which is primarily reflected by changes in the periodic components of the spectrum. A well described phenomenon is the slowing of the center frequency in the alpha range, that is integral to attention and cognition processes (Cesnaite et al., 2023).

Recent methodological advancements offer an apt means to decompose the EEG power spectrum into rhythmic oscillations (periodic component) and non-rhythmic fluctuations (aperiodic component). It helps to dissect complex neural signals (Leroy et al., 2022), enrich our understanding of brain function (Lendner et al., 2020), identify potential biomarkers of disease (Pollak et al., 2024), and assess the effectiveness of a therapy (Kundu et al., 2023; Salvatore et al., 2023). The strength of the approach particularly stems from its reflection of the EEG signal’s two-fold nature, encompassing both its mathematical characteristics in signal analysis and its neurophysiological correlates. The underlying aperiodic activity, distributed in a 1/f-manner, has been linked to the cortical balance of synaptic excitation and inhibition (Wang, 2020). In contrast, the superimposed periodic activity involves an interplay of cortical neural networks partly orchestrated by subcortical nodes (Seeber et al., 2019), and is estimated by gaussians centered around the oscillatory peaks.

Non-invasive brain stimulation techniques such as transcranial alternating stimulation (tACS) are increasingly used to entrain such oscillatory brain activity in elderly subjects and numerous neurological and psychiatric disorders, including but not limited to Parkinson’s disease (Madrid and Benninger, 2021), dementia (Manippa et al., 2023) and depression (Lee et al., 2022). The tACS is intended to interact with ongoing oscillatory activity, yet the frequency of these oscillations changes in the aging brain, resulting in possible mismatches (Fröhlich et al., 2015). When using the same fixed frequency in older and young individuals, this fact may limit the efficacy of the induced oscillatory electric fields. While the application of tACS using an individual peak frequency seems to be an ideal approach (Kasten et al., 2018; Vosskuhl et al., 2018), the identification of individual frequencies can be quite laborious and is not always feasible in the clinical setting. Furthermore, recent studies have shown that fixed stimulation protocols may even exhibit stronger aftereffects than individualized stimulation frequency (Ladenbauer et al., 2023; Stecher et al., 2021). To our opinion, this would however require an age-dependent adaptation of the applied oscillatory stimulation. It is currently unknown which exact “resonance” frequency should be chosen to interact with oscillations in a patient of a given age.

Therefore, this analysis of clinical data aims to describe the age-related periodic and aperiodic activity in 532 physiological resting-state EEG recordings by decomposing the spectrum into its periodic and aperiodic components. We derive a comprehensive method to estimate the age-adjusted center frequency within a band of interest. With this novel approach, we provide an alternative method to peak frequency determination to find the optimal age-adjusted stimulation frequency, that circumvents more laborious and impracticable methods to individualize the stimulation frequency.

## 2. Methods

This analysis was conducted with EEG data from a German tertiary care university hospital. This study adhered to the regulations of the local ethics committee. The quality standard regarding ethical and scientific data collection followed the ICH-GCP guidelines. Formal consent was obtained from the data protection board to allow handling and pseudonymization of clinical routine data.

### 2.1. Data collection

The analyzed EEG data was recorded between 2004 and 2014. Along with the EEG recording, a brief medical history as well as the interpretation of the neurologist in charge were assessed. The electrodes were placed with a common montage following the 10/20-system at the following 19 electrode positions: Fp1, Fp2, Fz, F3, F4, F7, F8, Cz, C3, C4, T7, T8, Pz, P3, P4, P7, P8, O1, O2. The EEG was recorded with a commercially available system used in the clinical routine (Galileo.NET, BE Light system, EB Neuro S.p.A., Firenze, Italy) with a sampling frequency of 256 Hz. The reference and ground electrode were placed at A1 and A2 respectively. Patients were asked to close their eyes during the recording, lasting around 20 minutes, interrupted by provocation maneuvers. Only recordings interpreted as “physiological EEG recording” were included in this analysis. We intentionally did not apply further exclusion criteria like age or medical conditions.

### 2.2. EEG Analysis

For each patient 20 epochs of 10 seconds free of artifacts or provocation maneuvers were selected. All analyses were conducted in MATLAB (MATLAB R2023b, 155 Natick, Massachusetts: The MathWorks Inc.; 2023.) with the Chronux toolbox (version 2.12 v03, http://chronux.org/) (Mitra and Bokil, 2008). Analyses were performed separately for every single patient, channel, and epoch. Preprocessing included trendline removal and bandpass filtering (0 – 45 Hz). The canonical frequency bands were defined as following: delta (δ, 1-4 Hz), theta (θ, 4-7 Hz), alpha (α, 7-12 Hz), low beta (β_1_, 12-20 Hz), high beta (β_2_, 20-30 Hz), and gamma (γ, 30-45 Hz).

A multitaper method was applied to estimate the power spectrum with a moving window length of 2 seconds, a shift of 0.1 seconds, a time-bandwidth product of 2 and 3 Slepian tapers. We did not conduct a normalization of the power spectrum to be able to assess age-related changes. The power spectrum was decomposed into its periodic and aperiodic components using the FOOOF toolbox with default settings: peak width limit 0.5–12 Hz, infinite maximum number of peaks, minimum peak height of 0 μV2, peak threshold of 2 standard deviations and a fixed aperiodic mode without a knee parameter (Donoghue et al., 2020).

We characterized the aperiodic offset and exponent, respectively the intercept and the slope of the aperiodic activity. The FOOOF toolbox parametrizes the periodic activity as fitted gaussians over the aperiodic slope with the center frequency, the adjusted periodic power, and the bandwidth. Instead of defining the power of the alpha peak as the total power within a canonical band, we searched if a power peak with a center frequency within the range of interest was fitted and the associated adjusted periodic power and bandwidth was assessed (Figure 1). If more than one power peak was comprised within the band of interest, the peak with the highest power was selected. Topographic representations were performed with the *topoplot* function in EEGlab (version 2023.1) (Delorme and Makeig, 2004).

**Figure 1:**
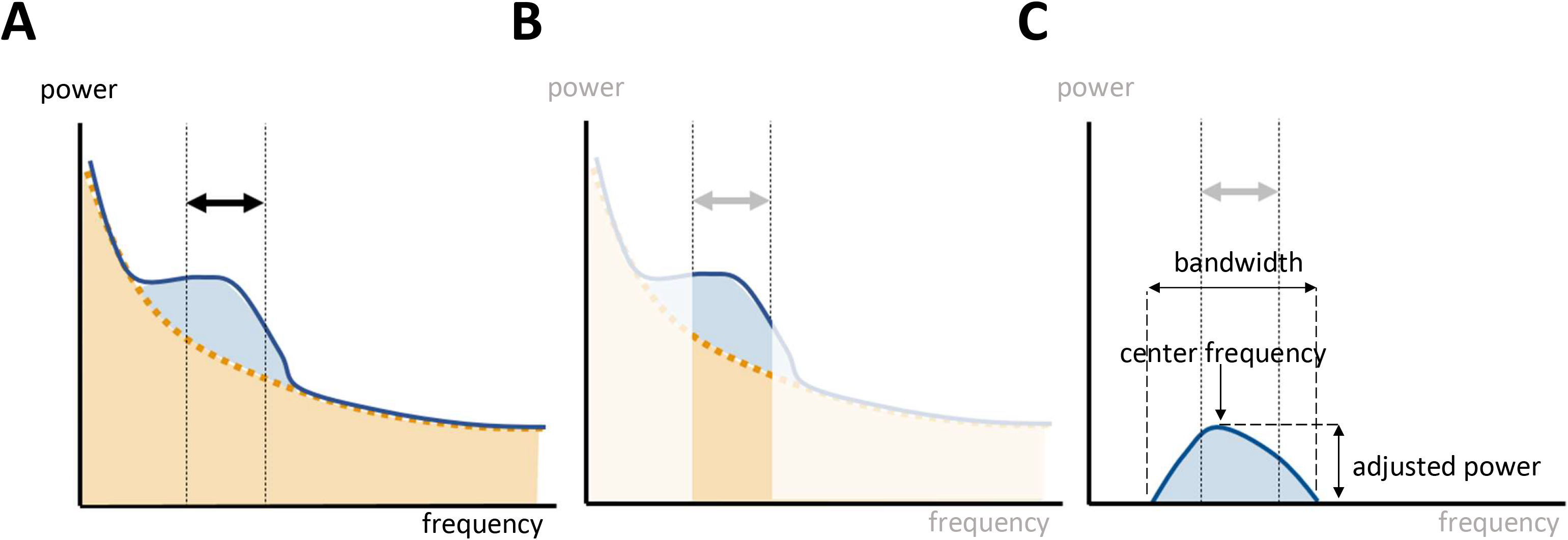
Parametrization of oscillatory activity within a frequency range of interest. **A**: Power spectrum decomposition with fitting of the aperiodic (orange) and periodic (blue) components. **B**: The power within a canonical band comprises the periodic and aperiodic components of the power spectrum. **C**: The FOOOF toolbox parametrizes the periodic activity by fitting gaussians of oscillatory activity over the underlying aperiodic slope.

### 2.3. Statistical Analysis

All statistical analyses were performed in MATLAB. Electrodes were grouped and averaged into five regions: frontal (Fp1, Fp2, Fz, F3, F4, F7, F8), central (Cz, C3, C4), temporal (T7, T8), parietal (Pz, P3, P4, P7, P8), and occipital (O1, O2). Linear regressions were fitted for each parameter with the *fitlm* function. We extracted the residual mean standard error (RMSE) – the spread of the empirical data around the linear fit –, the coefficient – the change in value each year –, and the intercept – the theoretical frequency at 0 years – for each parameter. The linear models were compared to models with only a constant to test for significance. Intrasubject variability of the peak center frequency was defined as the standard deviation of the center frequency within 20 epochs for each patient within a single EEG recording.

### 2.4. Calculation of Age-Adjusted Frequency

We defined the age-adjusted frequency based on the results of the linear regression models. The age-adjusted frequency can be calculated using with the linear formula:

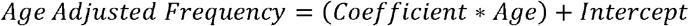

## 3. Results

A total of 10.620 EEG epochs from 532 patients were included in this analysis (Table 1 and Figure S1).

**Table 1:** Patient characteristics.

|  | Patient cohort |
| --- | --- |
| Sample size | 532 |
| Female (%) | 274 (52) |
| Age range (years) | 8-92 |
| Age mean (± standard deviation) | 50.5(±17.6) |

### 3.1. Center Frequency

An average topographical representation of the center frequency dynamics within all power bands can be found in Figure 2. A global age-related change in the center frequency was solely found in the alpha and gamma range. In the alpha band, there was a significant decrease of around 0.01 Hz per year, while we saw an increase in the gamma peak frequency between 0.01 and 0.03 Hz per year, depending on the region. In the other frequency bands, significant changes were regional and had smaller orders of magnitude. The RMSE depends on the frequency band and is generally larger in the faster oscillatory ranges. A similar dynamic was observed for intrasubject variability (Figure 2, Table S1).

**Figure 2:**
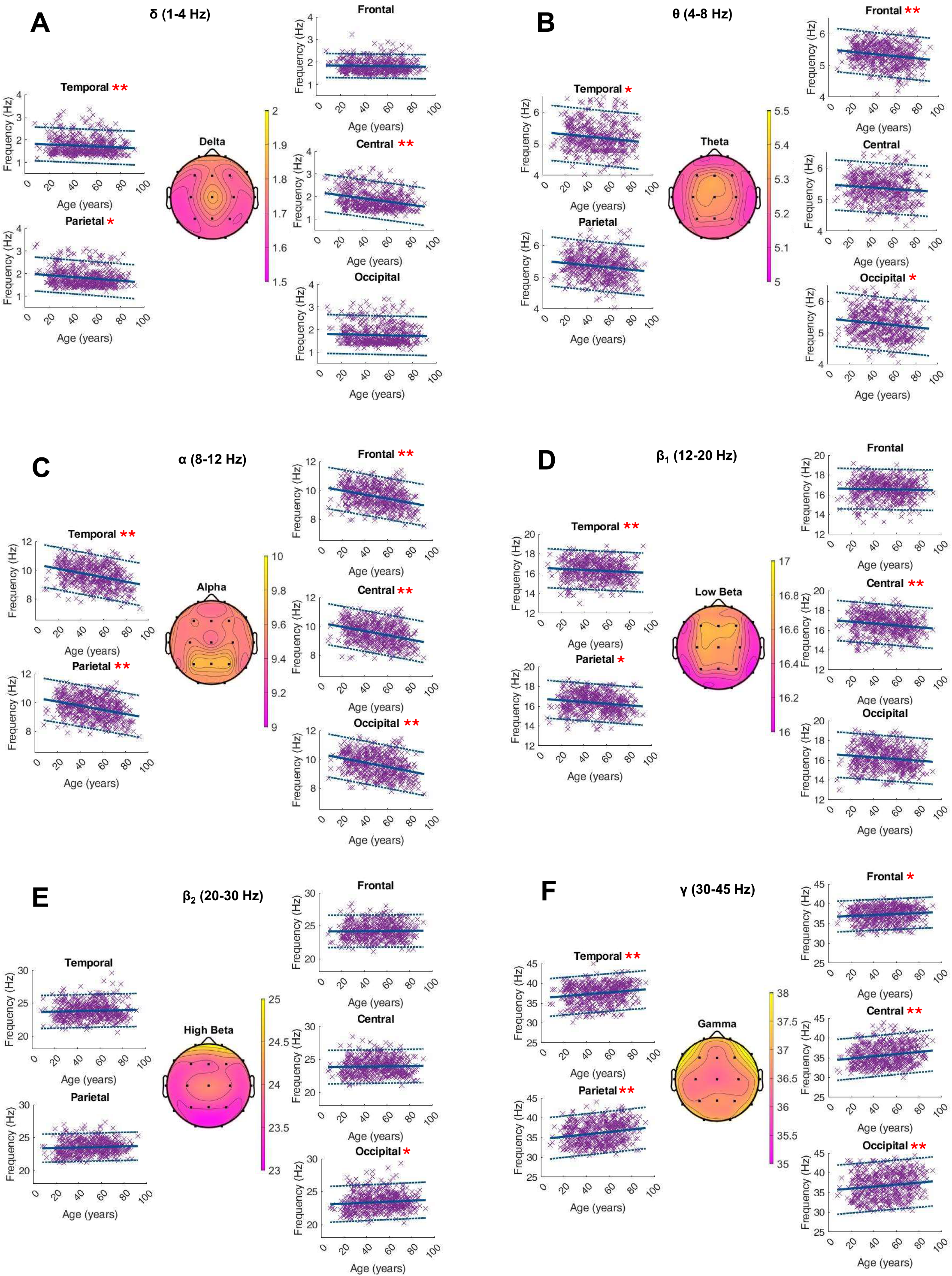
Characterization of peak frequency (Hz) within the frequency bands of interest on the average of 20 single 10-s epochs per patient. For each frequency band, an averaged topographical representation of the center frequency was computed, and the linear regression (thick blue line) as well as the 95% confidence interval (dashed blue lines) was plotted for each parameter. The frequency bands were defined as follows: delta (**A**, 1-4 Hz), theta (**B**, 4-8 Hz), alpha (**C**, 8-12 Hz), low beta (**D**, 12-20 Hz), high beta (**E**, 20-30 Hz), and gamma (**F**, 30-50 Hz). If the p-value was below the significance level of 0.05, the statistical significance was reported with red asterisks (*: p-value 0.05-0.001; **: p-value ≤0.001,).

### 3.2. Adjusted Peak Power

The age-related changes in the adjusted power of the fitted peaks is shown in Figure 3. Age did not show a globally significant effect on the adjusted power within any frequency band. The RMSE, coefficients, intercept, and p-values as well as the intrasubject variability of the adjusted power within the ranges of interest can be found in the supplementary materials (Figure 3, Table S2).

**Figure 3:**
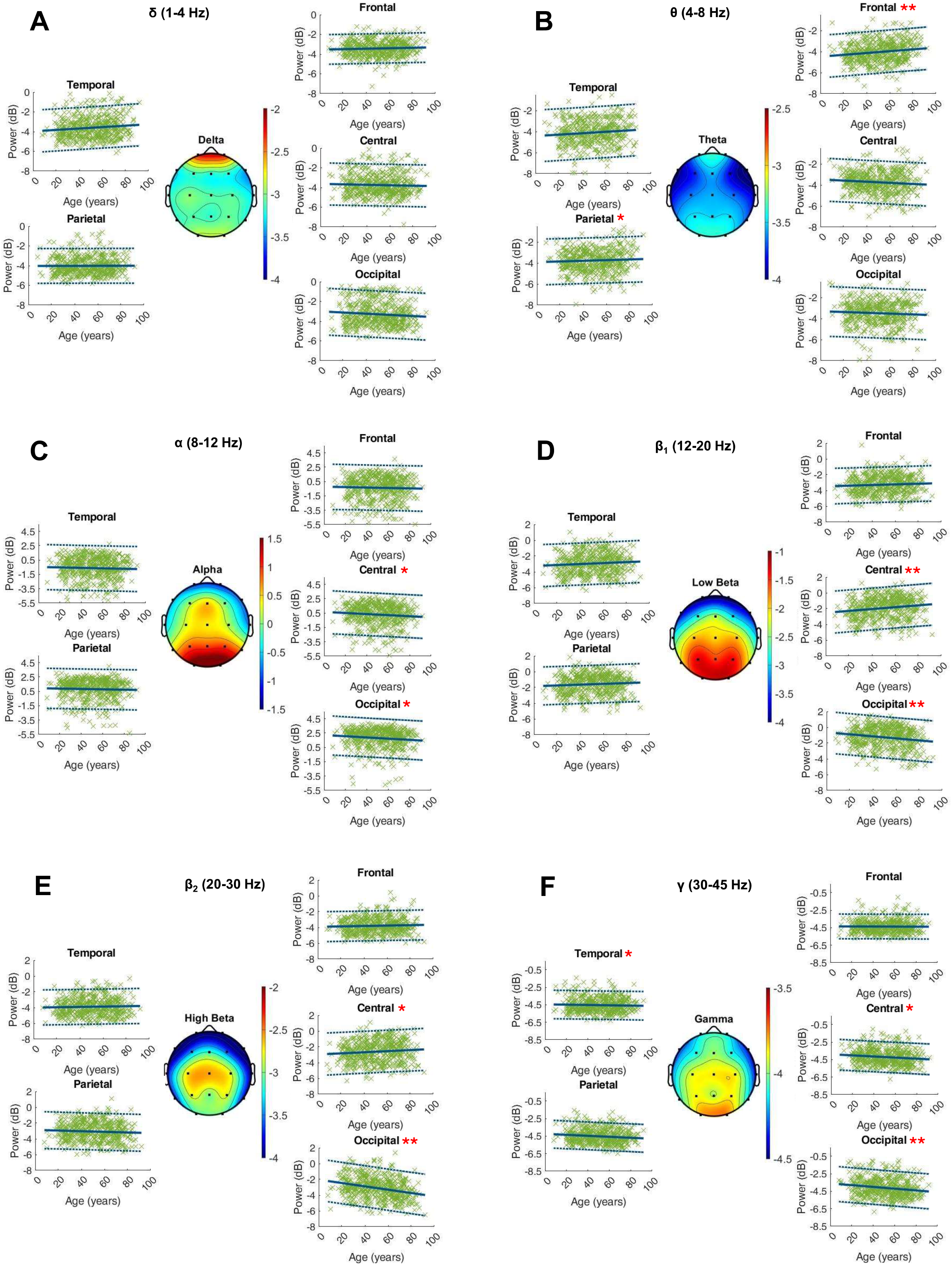
Characterization of peak power (dB) within the frequency bands of interest on the average of 20 single 10-s epochs per patient. For each frequency band, an averaged topographical representation of the peak power was computed, and the linear regression (thick blue line) as well as the 95% confidence interval (dashed blue lines) was plotted for each parameter. The frequency bands were defined as follows: delta (**A**, 1-4 Hz), theta (**B**, 4-8 Hz), alpha (**C**, 8-12 Hz), low beta (**D**, 12-20 Hz), high beta (**E**, 20-30 Hz), and gamma (**F**, 30-50 Hz). If the p-value was below the significance level of 0.05, the statistical significance was reported with red asterisks (*: p-value 0.05-0.001; **: p-value ≤0.001,).

### 3.3. Bandwidth

Age-related changes in the bandwidth of the fitted oscillatory peaks are depicted in Figure 4. There were no global dynamics for this parameter. In the low beta range, we saw a significant age-related increase of the peak bandwidth in the frontal, parietal, central and temporal electrodes of 0.01 to 0.02 Hz per year. The RMSE, coefficients, intercept, and p-values as well as the intrasubject variability of the peak bandwidth within the ranges of interest can be found in the supplementary materials (Figure 4, Table S3).

**Figure 4:**
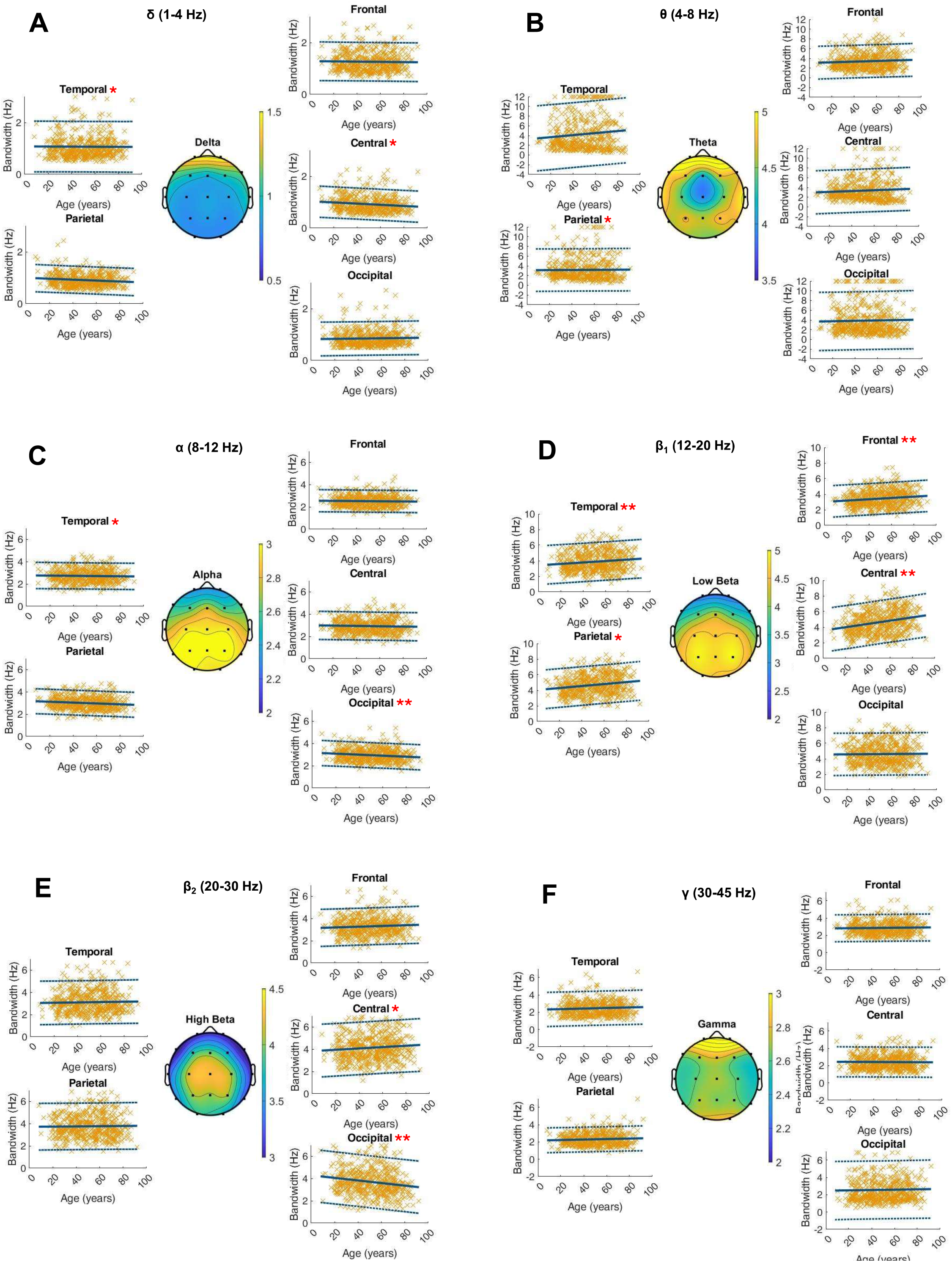
Characterization of peak bandwidth (Hz) within the frequency bands of interest on the average of 20 single 10-s epochs per patient. For each frequency band, an averaged topographical representation of the peak bandwidth was computed, and the linear regression (thick blue line) as well as the 95% confidence interval (dashed blue lines) was plotted for each parameter. The frequency bands were defined as follows: delta (**A**, 1-4 Hz), theta (**B**, 4-8 Hz), alpha (**C**, 8-12 Hz), low beta (**D**, 12-20 Hz), high beta (**E**, 20-30 Hz), and gamma (**F**, 30-50 Hz). If the p-value was below the significance level of 0.05, the statistical significance was reported with red asterisks (*: p-value 0.05-0.001; **: p-value ≤0.001,).

### 3.4. Aperiodic Activity

The aperiodic exponent showed a global significant decrease with age of around 0.003 μV^2^/Hz per year. We did not see age-dependent significant changes in the aperiodic offset (Table S4).

### 3.5. Example

We used this method to calculate the age-adjusted frequency in a tACS protocol to prevent the occurrence of postoperative delirium in elderly patients undergoing general anesthesia for major surgery (“https://drks.de/register/de/trial/DRKS00033703,” n.d.). In this tACS protocol applied on patients around 75 years of age, the age-adjusted frequency amounted to 9.5 Hz.

## 4. Discussion

This analysis provides the first spectrum-wide normative characterization of the age-related changes in oscillatory (periodic) and non-rhythmic (aperiodic) activity within physiological resting-state EEGs. While these processes have been partially described in some frequency bands or within age-cohorts, a parametrization of these dynamics over the life span was lacking. We found age-related changes in both periodic and aperiodic EEG parameters. Regarding the periodic activity, especially the center frequency of the alpha peak significantly decreased with age, while the gamma peak frequency increased. The aperiodic exponent describing the slope of the aperiodic activity decreased with age, but no significant changes were observed in the offset.

### 4.1. Normative Database of Oscillatory Activity

Quantitative EEG (qEEG) analysis emerged from digital signal analysis and spread itself quickly, due to increasing capacities of commonly used computers (Höller, 2021). In classical, clinical EEG analysis, the recordings are inspected visually by trained experts regarding the occurrence of specific patterns (Zhang et al., 2023). In contrast, qEEG analysis uses computer-based methods to breakdown EEG signals, allowing for the quantification of signal components both at specific channels and between channels (Gavaret et al., 2023). Normative databases of qEEG have been emerging and are essential to develop EEG biomarkers of diseases (Ko et al., 2021; Prichep, 2005). Here we present the first spectrum-wide normative database of periodic and aperiodic activity from a large cohort.

### 4.2. Age-Related Spectral Changes

We provided a normative characterization of ongoing oscillatory activity in the resting-state EEG in six frequency bands by isolating the periodic from the aperiodic components in the power spectrum. In line with previous studies, the aperiodic slope showed a ubiquitous decrease with age, significantly affecting the total power (Donoghue et al., 2020). Conventional decomposition of EEG spectra into canonical frequency bands does neither account for this, nor for frequency shifts across rigid band limits (Scally et al., 2018). For instance, we confirmed previous evidence that the major part of age-related differences in the alpha power can be explained by the flattening of the aperiodic slope and a shift towards the theta range, rather than a loss of oscillatory activity (Cesnaite et al., 2023; Merkin et al., 2023; Tröndle et al., 2023). Similarly, the loss of theta power associated with increasing age and deterioration of cognitive status can be explained from the flattening of the aperiodic slope (Caplan et al., 2015; Cesnaite et al., 2023).

### 4.3. Estimation of Age-Adjusted Center Frequency

The observed slowing in the alpha center frequency by approximately 0.01 Hz per year corresponds to previous findings and reinforces the notion that the individual alpha frequency (IAF) alters as the brain ages and cognition deteriorates (Cesnaite et al., 2023; Merkin et al., 2023). We observed an acceleration of the center frequency in the gamma range amounting to 0.01 to 0.03 Hz per year depending on the brain region. The acceleration of frequencies in the gamma range has been proposed as a compensatory mechanism to counteract for declining nerve conduction velocities (Hong and Rebec, 2012). It has been postulated that this mechanism contributes to the flattening of the aperiodic slope and is also reflected in the acceleration of the center frequency in the gamma oscillations (Voytek et al., 2015). For the other frequency bands, we show that there is no overall age-related trend in the center frequency and present normative, age-independent values.

### 4.4. Clinical Application

Recent work showed that tACS in the alpha range modulates the periodic but not the aperiodic components of the power spectrum, underlying the importance of this distinction in the context of brain stimulation (Kasten et al., 2024). tACS is a growing therapeutic field, yielding promising, yet inconsistent results (Herrmann et al., 2013). Neuroanatomical as well as neurophysiological variabilities may affect the individual (after-)effects of tACS (Zanto et al., 2021). To account for such variations, studies choose stimulation protocols that measure the IAF before the stimulation in the alpha range and use it as the frequency parameter of the stimulation. Yet, no consistent effects on spectral power modulation arises from stimulating at the IAF rather than a fixed frequency (De Koninck et al., 2023).

A possible reason for such inconclusive results may lie in the intrasubject variability of the alpha peak during a single recording, that amounted to 0.5 Hz within one resting-state EEG recording. This questions the rationale for a stimulation protocol that uses a specific IAF of each subject, as the determined IAF, even when measured immediately before a stimulation may still be inaccurate during the stimulation (Gray and Emmanouil, 2020; Haegens et al., 2014). An approach to correct for the dynamic changes in the IAF that synchronizes the stimulation to the concurrent EEG-activity may be a closed loop setup where the stimulation frequency is driven by the measured frequency in a simultaneous EEG recording (Nasr et al., 2022; Zrenner and Ziemann, 2024). However, this method is more laborious and limits the applicability in the clinical context.

These challenges motivated the here presented method of “individualization” through age group-dependent frequency adjustment, that provides a practical compromise between the more complex and error-prone closed-loop stimulation and the simple determination of stimulation frequency vaguely based on canonical frequency bands. It additionally ensures reproducibility across subjects and studies, contributing to a more standardized approach to tACS research that is critical to reveal the effects of the stimulation.

### 4.5. Limitations

A limitation of this analysis lies in its cross-sectional design, which does not allow for the tracking of individual aging processes over time. Consequently, longitudinal studies would offer a more nuanced understanding of how EEG parameters evolve with age in the same subjects. The study’s interpretations of frequency changes, especially in the gamma bands, may also be impacted by external variables such as minor physical movements or muscle artifacts, not accounted for during EEG recordings. Moreover, the analysis did not implement corrections for multiple testing since the primary goal was to describe physiological phenomena, rather than to validate or introduce new hypotheses.

## 5. Conclusions

The findings from this analysis of clinical routine data highlight the intricate dynamics of age-related changes in resting-state EEG signals, which may have crucial implications for the understanding of the aging brain and the tailoring of neurotherapeutic interventions.

## Data Availability

All data produced in the present study are available upon reasonable request to the authors

## Author Contributions

All authors developed the idea and concept for the methodological framework. SL & VB performed the data analyses and wrote the first draft of the manuscript. All authors reviewed and approved the final version of the manuscript.

