## Supplementary Files for "Normative characterization of age-related periodic and aperiodic activity in resting-state clinical EEG recordings"

### **Supplementary Material**

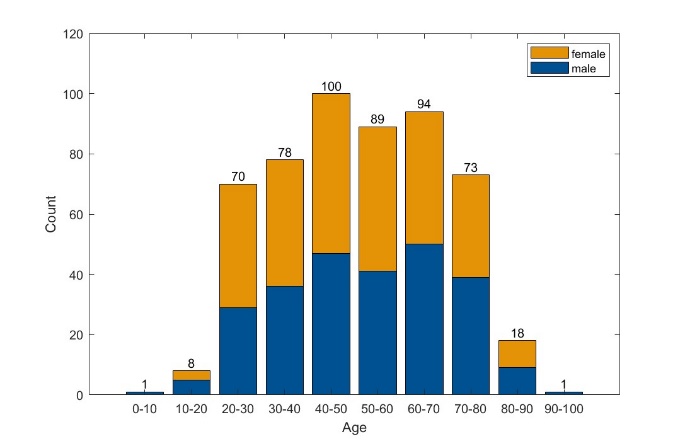

Figure S1: Histograms of age and sex distribution

Table S1: Coefficient, intercept, residual mean standard error (RMSE), and p-value of the fitted linear regression models for the peak center frequency (Hz) within bands of interest. Intrasubject variability was additionally computed over the 20 individual segments of each patient.

|  | Region | Coefficient | Intercept | RMSE | p-Value | Intrasubject Variability ± Standard Deviation |
| --- | --- | --- | --- | --- | --- | --- |
| Delta Peak Frequency | Frontal | -0.001 | 1.85 | 0.27 | 0.302 | 0.5±0.2 |
|  | Parietal | **-0.002** | **1.84** | 0.38 | **0.019** | 0.45±0.3 |
|  | Central | **-0.007** | **2.22** | 0.42 | **<0.001** | 0.51±0.3 |
|  | Temporal | **-0.004** | **2.01** | 0.38 | **<0.001** | 0.45±0.26 |
|  | Occipital | -0.001 | 1.81 | 0.44 | 0.282 | 0.48±0.36 |
| Theta Peak Frequency | Frontal | **-0.003** | **5.80** | **0.41** | **0.001** | 0.56±0.27 |
|  | Parietal | -0.001 | 5.56 | 0.59 | 0.513 | 0.47±0.37 |
|  | Central | -0.001 | 5.70 | 0.49 | 0.271 | 0.55±0.31 |
|  | Temporal | **-0.002** | **5.79** | 0.47 | **0.033** | 0.52±0.29 |
|  | Occipital | **-0.003** | **5.70** | 0.55 | **0.039** | 0.47±0.34 |
| Alpha Peak Frequency | Frontal | **-0.015** | **10.34** | 0.72 | **<0.001** | 0.56±0.27 |
|  | Parietal | **-0.015** | **10.45** | 0.74 | **<0.001** | 0.6±0.28 |
|  | Central | **-0.015** | **10.32** | 0.72 | **<0.001** | 0.57±0.28 |
|  | Temporal | **-0.015** | **10.39** | 0.74 | **<0.001** | 0.55±0.25 |
|  | Occipital | **-0.016** | **10.43** | 0.76 | **<0.001** | 0.47±0.26 |
| Low Beta Peak Frequency | Frontal | -0.002 | 16.64 | 1.04 | 0.447 | 1.88±0.46 |
|  | Parietal | **-0.005** | **16.58** | 1.00 | **0.034** | 1.92±0.44 |
|  | Central | **-0.010** | **17.05** | 1.03 | **<0.001** | 1.82±0.49 |
|  | Temporal | **-0.009** | **16.77** | 0.96 | **<0.001** | 1.83±0.43 |
|  | Occipital | **-0.009** | **16.63** | 1.16 | **0.003** | 1.83±0.55 |
| High Beta Peak Frequency | Frontal | 0.001 | 24.15 | 1.25 | 0.677 | 2.44±0.48 |
|  | Parietal | 0.004 | 23.60 | 1.27 | 0.238 | 2.48±0.63 |
|  | Central | 0.002 | 23.81 | 1.29 | 0.467 | 2.31±0.63 |
|  | Temporal | 0.004 | 23.35 | 1.08 | 0.139 | 2.38±0.54 |
|  | Occipital | **0.008** | **23.02** | 1.38 | **0.023** | 2.28±0.7 |
| Gamma Peak Frequency | Frontal | **0.013** | **36.66** | 1.97 | **0.010** | 4.03±0.71 |
|  | Parietal | **0.024** | **36.25** | 2.41 | **<0.001** | 4.05±1.04 |
|  | Central | **0.028** | **34.23** | 2.62 | **<0.001** | 4.19±1.27 |
|  | Temporal | **0.031** | **34.59** | 2.65 | **<0.001** | 4.14±1.19 |
|  | Occipital | **0.025** | **35.51** | 3.15 | **0.001** | 4.14±1.55 |

Table S2: Residual mean standard error, coefficient, intercept and p-value of the fitted linear regression models for the peak power (dB) within bands of interest. The intrasubject variability was additionally computed over the 20 individual segments of each patient.

|  | Region | Coefficient | Intercept | RMSE | p-Value | Intrasubject Variability ± Standard Deviation |
| --- | --- | --- | --- | --- | --- | --- |
| Delta Peak Power | Frontal | 0.0022 | -3.25 | 0.73 | 0.216 | 0.21±0.05 |
|  | Parietal | **0.0063** | **-3.55** | 1.03 | **0.013** | 0.21±0.08 |
|  | Central | -0.0028 | -3.23 | 1.04 | 0.271 | 0.2±0.08 |
|  | Temporal | -0.0008 | -3.62 | 0.89 | 0.711 | 0.2±0.07 |
|  | Occipital | -0.0053 | -2.70 | 1.10 | 0.053 | 0.19±0.1 |
| Theta Peak Power | Frontal | **0.0109** | **-4.23** | 1.03 | **<0.001** | 0.11±0.06 |
|  | Parietal | **0.0076** | **-4.24** | 1.23 | **0.015** | 0.09±0.07 |
|  | Central | -0.0036 | -3.26 | 1.11 | 0.197 | 0.11±0.07 |
|  | Temporal | 0.0038 | -3.57 | 1.14 | 0.180 | 0.1±0.07 |
|  | Occipital | -0.0042 | -2.94 | 1.19 | 0.161 | 0.1±0.08 |
| Alpha Peak Power | Frontal | -0.0027 | -0.05 | 1.60 | 0.490 | 0.25±0.08 |
|  | Parietal | -0.0034 | -0.28 | 1.57 | 0.388 | 0.23±0.07 |
|  | Central | **-0.0075** | **0.84** | 1.49 | **0.041** | 0.25±0.08 |
|  | Temporal | -0.0027 | 1.12 | 1.39 | 0.436 | 0.28±0.09 |
|  | Occipital | **-0.0080** | **2.39** | 1.35 | **0.016** | 0.32±0.13 |
| Low Beta Peak Power | Frontal | 0.0036 | -3.34 | 1.13 | 0.190 | 0.13±0.05 |
|  | Parietal | 0.0056 | -3.09 | 1.32 | 0.088 | 0.14±0.05 |
|  | Central | **0.0112** | **-2.38** | 1.32 | **0.001** | 0.16±0.05 |
|  | Temporal | 0.0048 | -1.74 | 1.19 | 0.106 | 0.17±0.05 |
|  | Occipital | **-0.0130** | **-0.50** | 1.30 | **<0.001** | 0.19±0.07 |
| High Beta Peak Power | Frontal | 0.0024 | -3.82 | 0.95 | 0.299 | 0.12±0.04 |
|  | Parietal | 0.0018 | -3.83 | 1.11 | 0.502 | 0.12±0.04 |
|  | Central | **0.0066** | **-2.84** | 1.33 | **0.045** | 0.14±0.04 |
|  | Temporal | -0.0046 | -2.73 | 1.17 | 0.108 | 0.14±0.05 |
|  | Occipital | **-0.0217** | **-1.85** | 1.33 | **<0.001** | 0.15±0.07 |
| Gamma Peak Power | Frontal | -0.0004 | -4.18 | 0.72 | 0.840 | 0.15±0.05 |
|  | Parietal | -0.0021 | -4.16 | 0.84 | 0.315 | 0.16±0.06 |
|  | Central | **-0.0061** | **-3.67** | 0.87 | **0.005** | 0.15±0.06 |
|  | Temporal | **-0.0056** | **-4.03** | 0.81 | **0.006** | 0.14±0.06 |
|  | Occipital | **-0.0095** | **-3.40** | 0.99 | **<0.001** | 0.14±0.07 |

Table S3: Residual mean standard error, coefficient, intercept and p-value of the fitted linear regression models for the peak bandwidth (Hz) within bands of interest. The intrasubject variability was additionally computed over the 20 individual segments of each patient.

|  | Region | Coefficient | Intercept | RMSE | p-Value | Intrasubject Variability ± Standard Deviation |
| --- | --- | --- | --- | --- | --- | --- |
| Delta Peak Bandwidth | Frontal | -0.0005 | 1.28 | 0.38 | 0.628 | 0.68±0.37 |
|  | Parietal | -0.0002 | 1.08 | 0.50 | 0.874 | 0.49±0.46 |
|  | Central | **-0.0023** | **1.03** | 0.30 | **0.002** | 0.35±0.32 |
|  | Temporal | **-0.0018** | **1.00** | 0.27 | **0.007** | 0.34±0.26 |
|  | Occipital | 0.0006 | 0.82 | 0.33 | 0.490 | 0.3±0.37 |
| Theta Peak Bandwidth | Frontal | 0.0063 | 3.02 | 1.52 | 0.094 | 2.83±2.01 |
|  | Parietal | **0.0172** | **3.44** | 3.25 | **0.038** | 2.44±2.41 |
|  | Central | 0.0067 | 3.04 | 2.08 | 0.194 | 2.57±2.15 |
|  | Temporal | -0.0006 | 3.14 | 1.84 | 0.891 | 2.63±2.21 |
|  | Occipital | 0.0066 | 3.60 | 2.88 | 0.363 | 2.5±2.33 |
| Alpha Peak Bandwidth | Frontal | -0.0008 | 2.49 | 0.49 | 0.529 | 0.97±0.62 |
|  | Parietal | -0.0008 | 2.71 | 0.58 | 0.572 | 1.02±0.62 |
|  | Central | -0.0013 | 2.94 | 0.64 | 0.405 | 1.08±0.58 |
|  | Temporal | **-0.0037** | **3.13** | 0.56 | **0.008** | 1.02±0.54 |
|  | Occipital | **-0.0045** | **3.13** | 0.57 | **0.001** | 0.93±0.57 |
| Low Beta Peak Bandwidth | Frontal | **0.0084** | **3.02** | 1.02 | **0.001** | 2.03±0.83 |
|  | Parietal | **0.0093** | **3.41** | 1.25 | **0.003** | 2.34±0.81 |
|  | Central | **0.0212** | **3.56** | 1.41 | **<0.001** | 2.72±0.78 |
|  | Temporal | **0.0128** | **4.03** | 1.26 | **<0.001** | 2.7±0.67 |
|  | Occipital | 0.0010 | 4.55 | 1.37 | 0.766 | 2.73±0.83 |
| High Beta Peak Bandwidth | Frontal | 0.0033 | 3.13 | 0.84 | 0.112 | 2.04±0.78 |
|  | Parietal | 0.0015 | 3.04 | 0.99 | 0.530 | 2.02±0.92 |
|  | Central | **0.0060** | **3.83** | 1.20 | **0.042** | 2.5±0.8 |
|  | Temporal | 0.0009 | 3.72 | 1.06 | 0.728 | 2.36±0.83 |
|  | Occipital | **-0.0116** | **4.29** | 1.19 | **<0.001** | 2.31±0.98 |
| Gamma Peak Bandwidth | Frontal | 0.0012 | 2.80 | 0.79 | 0.532 | 2.39±1.16 |
|  | Parietal | 0.0032 | 2.29 | 1.00 | 0.192 | 2.07±1.45 |
|  | Central | -0.0007 | 2.44 | 0.88 | 0.758 | 1.98±1.62 |
|  | Temporal | 0.0028 | 2.17 | 0.73 | 0.119 | 2±1.65 |
|  | Occipital | 0.0020 | 2.46 | 1.70 | 0.636 | 1.91±1.84 |

Table S4: Coefficient, intercept, residual mean standard error (RMSE), and p-value of the fitted linear regression models for the aperiodic exponent (µV^2^/Hz) and offset (µV^2^).

|  | Region | Coefficient | Intercept | RMSE | p-Value |
| --- | --- | --- | --- | --- | --- |
| Exponent | Frontal | **-0.003** | **1.55** | **0.23** | **<0.001** |
|  | Parietal | **-0.004** | **1.34** | **0.29** | **<0.001** |
|  | Central | **-0.003** | **1.49** | **0.20** | **<0.001** |
|  | Temporal | **-0.003** | **1.47** | **0.20** | **<0.001** |
|  | Occipital | **-0.003** | **1.57** | **0.22** | **<0.001** |
| Offset | Frontal | 0.003 | -4.24 | 3.76 | 0.73 |
|  | Parietal | 0.004 | -4.73 | 3.69 | 0.69 |
|  | Central | 0.004 | -4.71 | 3.68 | 0.68 |
|  | Temporal | 0.004 | -4.61 | 3.70 | 0.68 |
|  | Occipital | 0.002 | -4.29 | 3.74 | 0.86 |
